# Reconsidering Brain Age: Why Age-Prediction Models Fail as Measures of Brain Aging

**DOI:** 10.64898/2026.05.07.26352620

**Authors:** Edvard O.S. Grødem, Stephen M. Smith, Didac Vidal-Piñeiro, Maxwell L. Elliott, the Alzheimer’s Disease Neuroimaging Initiative, Kristine B. Walhovd, Anders M. Fjell

## Abstract

Brain age models - machine-learning predictions of chronological age from brain imaging - are widely interpreted as markers of accelerated brain aging. Here we show that this interpretation cannot be supported. Because these models are trained to predict chronological age, they prioritize features that change similarly across people and actively downweight features that capture differences in individual trajectories, precisely the property an aging-rate biomarker must have. In effect, brain age models are optimized to ignore the very signal they are used to study, thereby risking converting stable between-person differences into apparent accelerated aging. Using theoretical analysis, simulations, and longitudinal MRI, we confirm both predicted failure modes: brain age models indicated “accelerated aging” in participants with low birth weight despite no longitudinal evidence, while a single hippocampal volume measurement was more sensitive than the brain age gap to tau-related neurodegeneration. Across much of the brain age literature, it is therefore not possible to determine whether reported effects reflect brain aging or stable anatomical differences, and the brain age gap should not be interpreted as a marker of brain aging or brain health. We propose alternative strategies that reorient prediction targets from shared age-related patterns to individual differences in change.

## 1 Introduction

Machine-learning–derived brain age has become a widely used imaging-based summary measure in human neuroscience (Fig 1), with the difference between predicted and chronological age - the “brain age gap” - commonly interpreted as reflecting accelerated or decelerated brain aging [1–4]. This interpretation is appealing because it appears to provide a single, intuitive marker of individual aging from cross-sectional data, potentially bypassing the need for longitudinal observation. Accordingly, brain age models have been applied across a wide range of domains, including schizophrenia [5–8], depression [9] and other brain disorders [10], as a function of genetic risks [11– 14], lifestyle and health-related factors [15–20], prenatal conditions [21], female health [22, 23], geographic diversity [24], mortality risk [25], pollution exposure [4] and even brain development [26, 27], and has been suggested as a potential outcome measure for prevention and intervention studies [28].

**Fig. 1.**
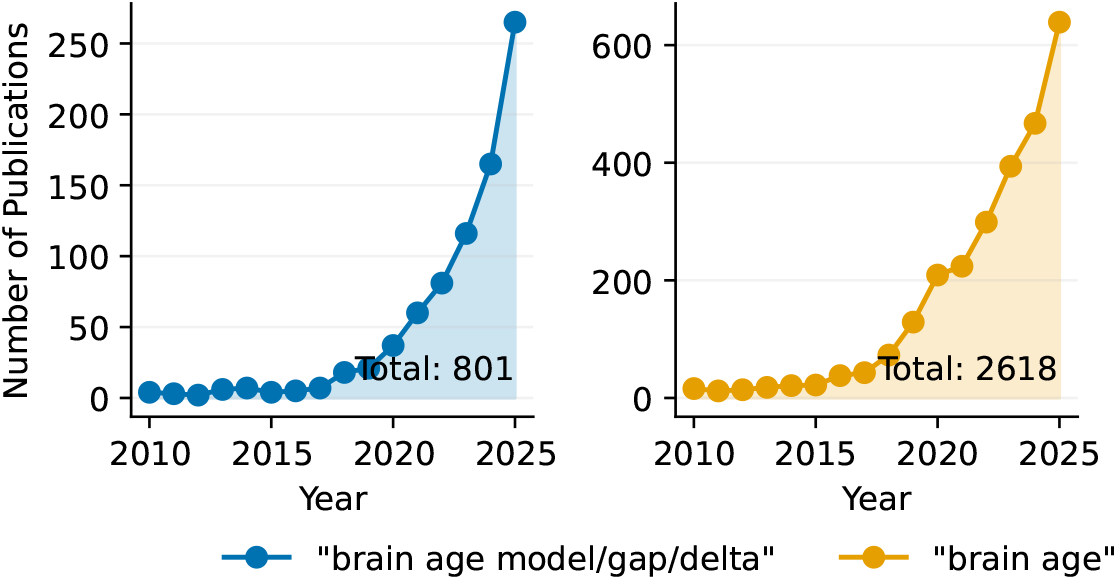
Growth in brain age publications. Left: number of publications per year identified in OpenAlex using terms related to “brain age gap/delta” or “brain age model” in the title or abstract. Right: number of publications per year identified using “brain age” in the title or abstract.

However, this interpretation rests on an assumption that has rarely been examined: that a model trained to predict chronological age is sensitive to individual differences in rates of brain change. Here we argue that the opposite is in fact true. The objective that defines brain age models, accurate prediction of chronological age, systematically prioritizes features that vary with age in a similar way across individuals, while down-weighting features that capture how individuals differ in their trajectories over time. Yet, it is precisely these differences in trajectories that define variation in aging rate. This creates a fundamental mismatch between what brain age models are optimized to detect and what they are typically assumed to measure. A biomarker of aging rate must be sensitive to between-person differences in longitudinal change. Brain age models, by design, suppress exactly this signal, but do not fully eliminate stable between-person differences in brain structure, particularly when these differences correlate with chronological age. As a consequence, lifelong anatomical differences that correlate with chronological age can be misclassified as apparent “accelerated” or “decelerated” aging, while genuine differences in ongoing neurobiological change are attenuated. This is not a limitation that can be resolved by larger samples, improved preprocessing, or more advanced machine learning algorithms, because it follows directly from the learning objective itself. Accordingly, the brain age gap can not be interpreted as a measure of individual differences in brain aging without additional evidence. In effect, the brain age objective is blind to the variation it is intended to measure.

Empirical findings have begun to reveal inconsistencies with brain age models that are difficult to reconcile with the standard interpretation. Several studies report that brain age gap correlates poorly, if at all, with longitudinal measures of brain change [29–31], calling its validity as an index of brain aging into question. At the same time, recent work shows that most of the variance in many brain features, at least up through middle age, can be explained by stable individual differences [32, 33], consistent with findings that brain age gap relates more strongly to early-life factors such as birth weight than to subsequent brain atrophy [29]. A UK Biobank study further showed that variance in brain age gap in middle- and older-aged adults is explained roughly three times more strongly by fixed baseline differences than by variation in aging rate [33]. This pattern aligns with longstanding evidence that individual differences in brain structure are established early in life and account for a substantial proportion of inter-individual variance across the lifespan [34–36].

This argument yields a clear empirical prediction: if brain age models are structurally misaligned with the goal of measuring aging, they should produce both false-positive and false-negative inferences about individual differences in brain aging. A valid biomarker of aging must be sensitive to between-person differences in longitudinal change [37, 38], and failure to capture this signal should lead directly to systematic misclassification and reduced sensitivity to genuine aging-related variation. To address this, we combine (i) a formal analysis of the brain age learning objective, (ii) controlled simulation experiments in which level and slope effects can be isolated, and (iii) empirical tests in longitudinal neuroimaging data designed to produce complementary failure cases.

For testing in real data, we construct two complementary failure tests derived from the theoretical framework. In the first scenario, a false-positive test, we examine birth weight, which is characterized by stable and lifelong positive associations with brain volume with little or no evidence for differences in aging rates [29, 34, 36]. Any association between birth weight and brain age gap is therefore unlikely to reflect differences in aging rate, but rather arise from misclassification of level effects. In the second scenario, a false-negative test, we use tau accumulation measured by PET, a biomarker associated with accelerated neurodegeneration characterized by differences in longitudinal change rather than baseline volume [39]. Based on the theoretical argument, we predict that brain age models will (i) produce apparent evidence of accelerated aging in the first scenario despite the absence of longitudinal differences, and (ii) show reduced sensitivity to tau-related neurodegeneration in the second scenario relative to simple volumetric measures. In other words, brain age models are expected to produce both false positives and false negatives with respect to individual differences in brain aging. Together, these analyses test a central implication of the argument: whether models trained to predict chronological age can recover individual differences in brain aging from cross-sectional data. If the reasoning above is correct, they will systematically misclassify stable between-person differences as aging effects while remaining relatively insensitive to genuine longitudinal change.

## 2 Results

### 2.1 The brain age objective suppresses signals of individual aging

Here we summarize our findings of the analysis in a non-technical narrative language for the general reader, proving that, under general assumptions, models trained to predict chronological age suppress the signals of individual differences in aging rates (see the Appendix for derivations).

Brain age models rely on a multivariate set of brain-derived features. In a cohort followed across adulthood, the distribution of features at each age can be summarized by an age-dependent mean and an age-dependent covariance. The covariance at a given age consists of three sources of variance:

- Measurement noise: non-persistent fluctuations (e.g., hourly/weekly fluctuations, scanner noise, and preprocessing variability).
- Baseline differences (level effect): stable inter-individual differences present at baseline that propagate over time (e.g., neuroanatomical size differences part of the constitution at birth, i.e. genetic differences and prenatal brain development).
- Individual differences in change (slope effects): deviations from the baseline trajectory, accumulating over time depending on, e.g., exposures, diseases, and resilience factors.

The third component is the key: it captures participants drifting away from the normative trajectory as they age; aka the slope effects. If these deviations align with the normative direction of aging, they are naturally interpreted as accelerated aging.

Most brain age models are trained to minimize error in predicting chronological age from the brain-derived features, typically the mean squared error. Regardless of whether the model is linear (e.g., Lasso or Ridge penalised regression) or highly non-linear (e.g., Random Forest, Neural Network), the optimal solution to this objective prioritizes features that offer the best signal-to-noise ratio for age prediction. Concretely, the model places a large weight on features where the mean changes reliably with age and downweights features that are highly variable across individuals of the same age. *Since variance from individual differences in change contributes to the total variance of a feature, this signal will actively be suppressed by the brain age objective*. Ironically, this is the signal most researchers studying aging care about. Depending on the type of model used, we find that the behavior is slightly different. Here we investigate two types: (i) a linear model and (ii) an optimal non-linear model. By optimal, we mean that given the true distribution of the data, the model infers the expected age. In a linear model, the same coefficients are used across age. The coefficients are selected by considering how age-dependent a feature is compared to the mean variance of that feature across all ages. We have previously demonstrated that it takes some time before inter-individual differences in aging accumulate enough to influence the total variance of the features [32]. Thus, the variance from individual differences in change will only partially affect the coefficients of the linear brain age model, since the variance is pooled across a wide age-range. Therefore, a linear brain age model might detect some inter-individual differences in aging, ironically partly due to a large part of the data used to fit the model not having much individual difference in aging. However, this sensitivity is much less than the potential sensitivity of the same features due to the suppressive nature of brain age models.

For a nonlinear model, we find that the suppressive effect of a brain age model can be even more severe. Non-linear brain age models can be highly accurate for classifying chronological age, especially those that are based on convolutional neural networks (CNN) [40], with a reported mean absolute error as low as 2 years, much less than the full age range (often 50-90 years). By linearizing the model around a chronological age we can investigate how the different features are used. If the model is un-biased, the expected brain age is the same as the chronological age. In this local view, we show that the model coefficients are determined by two quantities: (i) they are proportionally related to how quickly the feature mean changes with age, and (ii) they are inversely proportional to the total variance of the feature at that age. We find that this means that if the variance of a feature is dominated by individual differences in longitudinal aging changes, it will be nearly perfectly suppressed by a non-linear model at that age.

The variance from baseline is also suppressed in a brain age model. Hence, brain age models are optimized for features with little spread and high age dependency. Thus, brain age models should be less sensitive to baseline effects; however, since the baseline effects are usually much larger than the slope effects [32], these baseline effects will still highly influence the brain age model.

In summary, our model evaluation shows that the brain-age learning objective rewards features with strong cross-sectional age correlations - whether these arise from changes that track age consistently across individuals or from cohort effects that merely resemble aging. The same objective penalizes features whose age-related change varies between individuals. Yet, this between-person variability is precisely what one would need to capture in order to quantify differential aging.

#### 2.1.1 Simulated model experiment: A brain age model with two features

To demonstrate how brain age models are suppressing individual differences in aging rates, we create a small simulation experiment. We include two features, F1 (e.g., grey matter volume) and F2 (e.g., ventricular volume), shown in Fig. 2. At young age, we set their slope and variance to be the same; however, we define F1 to have no individual differences in the change rate, while F2 gradually accumulates differences. We see that this causes F2 to be much more spread out than F1 at old age.

**Fig. 2.**
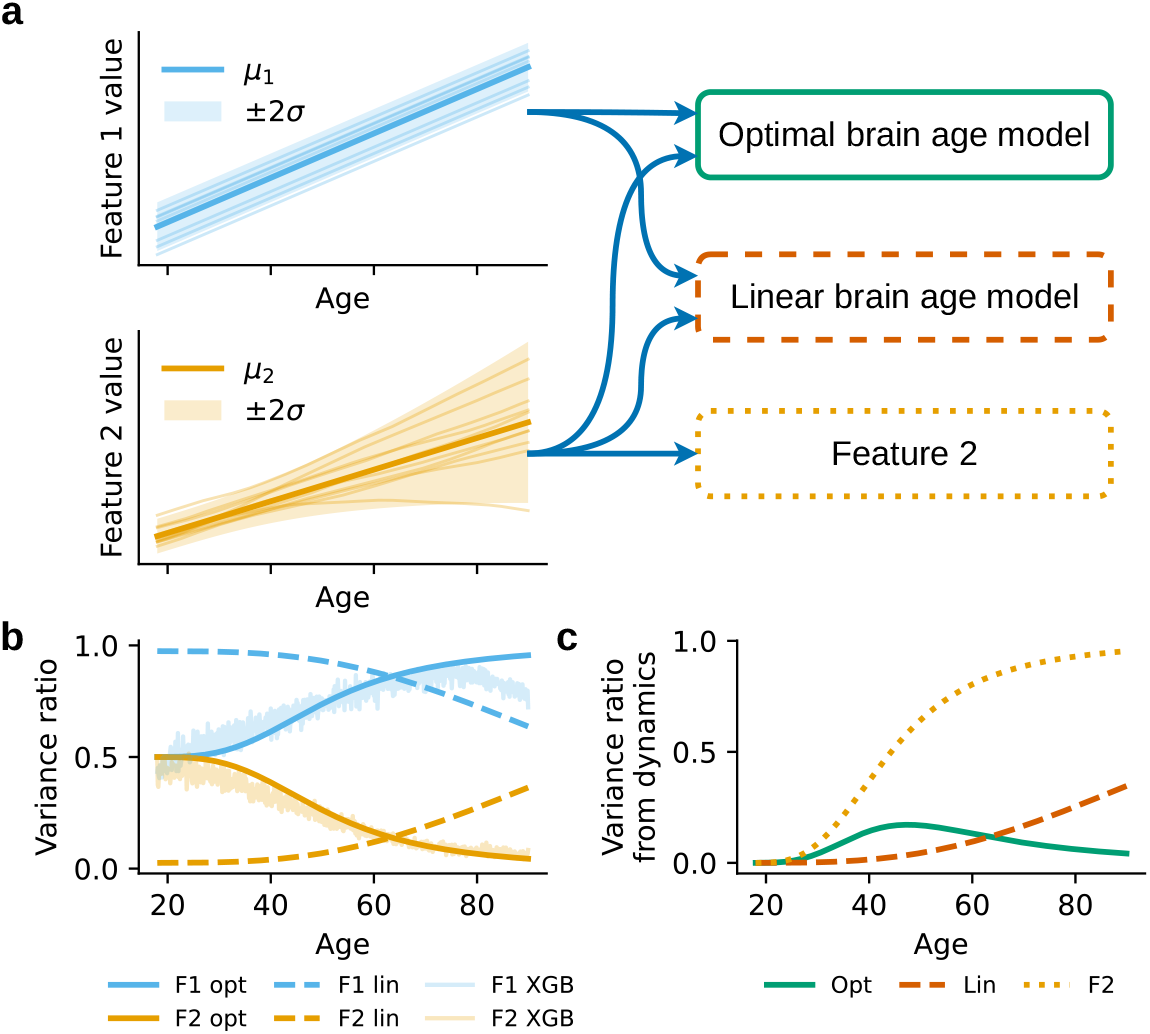
Feature contribution to brain age. In this simulation we show the suppressive nature of brain age models when analyzing accelerated aging. **a:** Two features are simulated. Feature 1 (F1), is highly age dependent, but after an initial offset, everyone changes at the same rate throughout life. Feature 2 (F2), is also highly age dependent, but with individual differences in change. After an initial offset, some people are changing faster, while others are changing slower, causing a spread of the distribution at older age (delta expansion). Three models are used to analyze the features: a perfect brain age model given the features, a linear brain age model, and just using F2 directly. **b:** The relative contribution of each feature to the brain age models at each chronological age. An XGBoost (XGB) model is also fitted to the simulated data with the linearized contribution of each feature to this model shown. **c:** The explained variance of the individual trajectories to the model outputs over the lifespan.

We use three models to “analyze” these data: (1) An optimal non-linear brain age model giving us the expected age given the two features, *E*[*t*|*F*1, *F*2], (2) a linear brain age model, and (3) a model that only uses F2 directly. We also fit an XGBoost model to the data to show that our theoretical predictions match practical brain age models. In this synthetic experiment, we know the generative model of the features, and we can thus compute the contributions of F1 and F2 to the models, which is shown in Fig. 2**b**. For the optimal brain age model, the two features start with equal contributions to the estimate of brain age, since they have the same variance and the same slope. However, we find that as F2 increases in variance, its contribution shrinks to almost zero. We see that the XGBoost model [41] matches the behavior of our theoretical predictions. We then compute the explained variance of the accumulated individual differences in aging to the model output shown in Fig. 2**c**. Here, we find that F2 at high age is dominated by accumulated individual differences in age change. The linear model does capture some of the individual differences due to the same weight being used over the whole lifespan. Interestingly, we find that the optimal brain age model captures almost none of the individual differences in aging at old age, due to the suppression of F2.

### 2.2 Empirical experiment: Testing brain age model performance in a double dissociation scenario

**Table 1.** Summary of the sample included in the empirical experiment. N is the total number of subjects used in the analysis, and N long is the number of subjects with longitudinal MRI. N Est. Tau is the number of subjects with at least one PET tau scan.

| Dataset | Name | Citation | N | N Long | M/F | Age Mean | Age SD | N Birth Weight | N Est. Tau |
| --- | --- | --- | --- | --- | --- | --- | --- | --- | --- |
| ADNI | Alzheimer's Disease Neuroimaging Initiative | [42] | 762 | 433 | 0.51 | 72.59 | 6.83 | 0 | 762 |
| DLBS | Dallas Lifespan Brain Study | [43] | 131 | 125 | 0.60 | 66.48 | 9.24 | 0 | 131 |
| HABS | Harvard Aging Brain Study | [44] | 195 | 187 | 0.59 | 75.49 | 6.14 | 0 | 195 |
| PreventAD | Pre-symptomatic Evaluation of Experimental or Novel Treatments for AD | [45] | 111 | 111 | 0.74 | 64.58 | 4.87 | 0 | 111 |
| LCBC | Center for Lifespan Changes in Brain and Cognition cohort | [46] | 160 | 108 | 0.68 | 62.11 | 7.82 | 160 | 0 |
| UKB | UK Biobank | [47] | 42319 | 2822 | 0.53 | 65.03 | 7.47 | 26535 | 0 |

Two scenarios are constructed as complementary failure tests, derived directly from our theoretical analysis. In the first scenario, a false-positive test, we use birth weight as the predictor variable. Birth weight is fixed at birth and therefore cannot itself change with age, and prior work establishes that higher birth weights are associated with greater brain volume throughout life but no difference in rate of volume change. Any signal a brain age model produces in association with birth weight therefore is unlikely to reflect differential aging, but rather a level effect being misread as accelerated or decelerated aging. Our analysis predicts precisely this misclassification: the model will detect a birth weight association and interpret it as a difference in brain age, when no aging-related signal exists.

In the second scenario, a false-negative test, we use accumulation of tau as measured by PET. Tau is a hallmark biomarker of late-life neurodegenerative disease, and brain age models are widely applied to detect tau- and Alzheimer’s-related change. Tau is undetectable early in life, so any signal reflects accumulation. Prior work links tau accumulation to faster volume loss but not to differences in initial volume [39]. Tau therefore represents the kind of signal brain age models should be sensitive to, i.e., genuine, individually-varying age-related change. Our analysis predicts the model will be less sensitive to tau-related change than a simple measure of hippocampal volume, which is not penalized for reflecting between-person variability in change. Together, these scenarios test whether brain age models behave as our theoretical analysis predicts when applied to real data: misclassifying level effects as spurious aging signals while remaining relatively insensitive to the very between-person variability in change that differential aging is meant to describe. We used structural MRI data from multiple cohorts (ADNI, DLBS, HABS, PreventAD, LCBC, and UK Biobank, see Tab. 1), processed using FreeSurfer to extract measures of volume, thickness and area, averaged across hemispheres to avoid collinearity resulting in 121 features. All imaging features were corrected for site effects using generalized additive models while accounting for age. The top and bottom 0.1% features were clipped to remove outliers. A detailed method description is provided in the Supplementary Information.

Brain age prediction models were trained on cross-sectional data from UK Biobank (UKB, n = 42,590). An Elastic Net regression model and a non-linear XGBoost model were trained to predict chronological age. Hyperparameters were optimized using grid search on the validation subset. Most of the birthweight data is in the UKB sample and to take advantage of the full dataset, five-fold cross validation was used, where 3 folds were used for training data, one for validation, and one for testing. By rotating which fold was used as the test fold, we were able to acquire a brain age estimate for all individuals without data-leakage. On the other datasets, an ensemble of the UKB trained brain age models was used. The top 5 features across the Elastic Net models are shown in Fig. 3**c**.

**Fig. 3.**
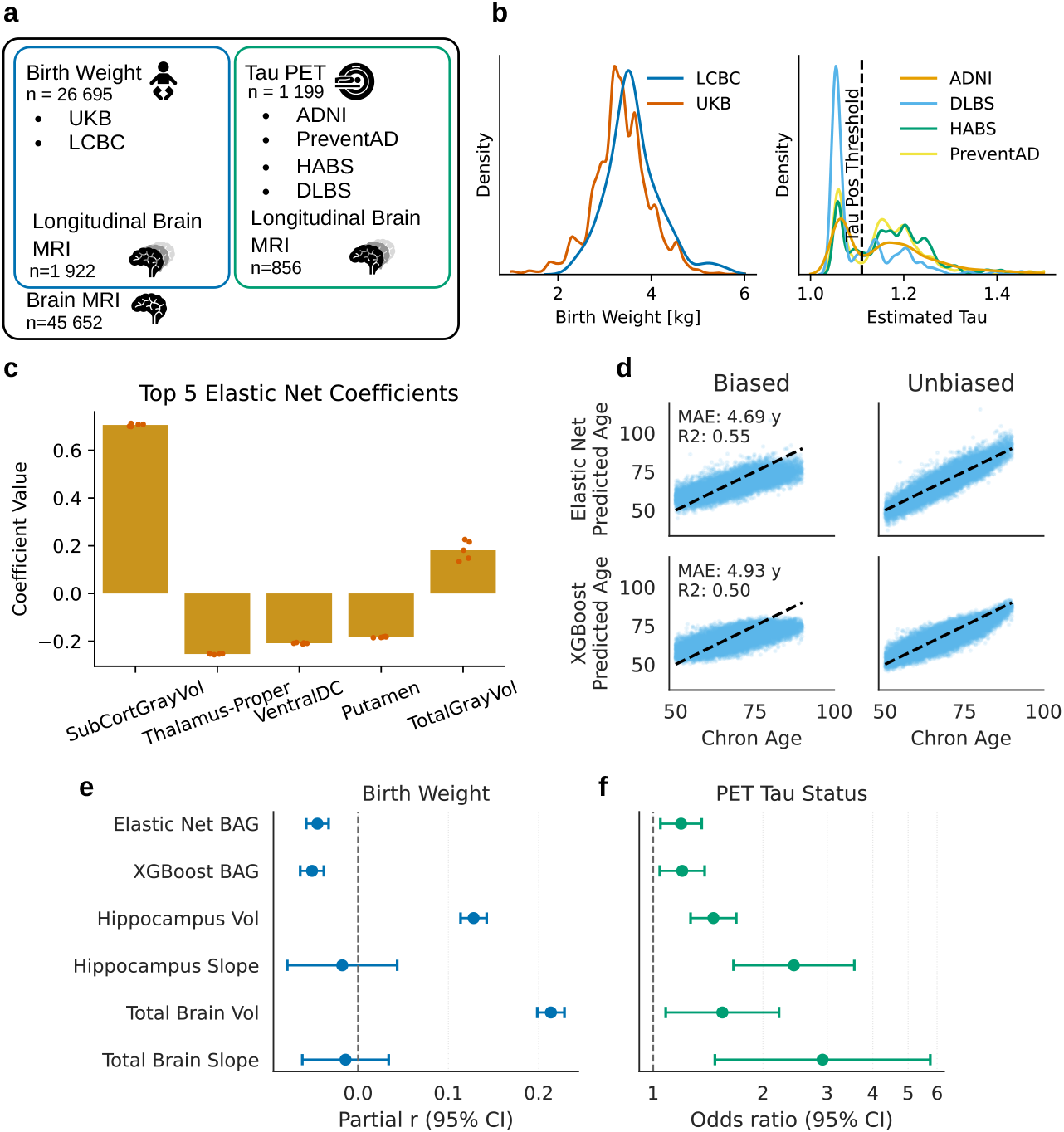
Testing brain age model performance in a double dissociation scenario. **a**: Samples used in this analysis. **b**: Distribution of birth weight and estimated tau PET values in the sample. **c**: The top 5 coefficients of the Elastic Net models. The coefficients are normalized such that their values squared sum to one. The red dots show the coefficients from each training fold in UKB, while the bars show the mean coefficient. **d**: Brain age prediction. The outputs of the brain age models are unbiased such that for each age the brain age gap is centered around the chronological age. **e**: The partial correlation (partial r) relating brain age gap (BAG) and volumetric measures to birth weight. A linear model is fitted to the data with correction for age, sex and imaging site. **f** : The odds ratio relating the brain age gap and volumetric measures to tau positivity from PET scans. A logistic regression model is used correcting for age, sex, imaging site and ICV. For visualization, a log scale is used and the signs of the volumetric measures are flipped such that a lower volume or slope corresponds to a higher risk of PET positivity.

Because brain age predictions typically show a systematic age-dependent bias [48], predicted ages were corrected using a Gaussian kernel approach. Specifically, the mean predicted age within a Gaussian window (standard deviation = 2 years) centered on each chronological age was estimated in the training data and subtracted from the predictions. This procedure removes both linear and non-linear age bias as shown in Fig 3**d**. The brain age gap was then computed as the difference between the bias-corrected predicted age and chronological age.

For comparison with simpler structural measures, we selected hippocampal and total brain volume and the longitudinal slope of each for each participant. The top and bottom 1% of the slope estimates were clipped to remove outliers. For cross-sectional analyses, one session per participant was selected as the session closest to the participant’s mean age across visits, which reduces regression-to-the-mean effects when slope estimates are included.

#### Dissociation 1: Stable individual differences in level but not slope

We first tested birth weight in relation to brain age gap using the Elastic Net and the XGBoost model outputs (n = 26,695). Both the covariates and the outcome were z-scored and we controlled for age using a smooth spline with 5 degrees of freedom, as well as a categorical variable for imaging site. Since the precision of the slope estimate is linearly dependent on the time interval between the first and last session squared [32], we weighted the volumetric slopes using the total observation time of each individual. This ensures that slope estimates derived from longer observation windows contributed proportionally more to the regression estimates. The results are presented in Fig. 3**e**. As predicted from the theoretical arguments, the brain age models indicated that lower birth weight was associated with “accelerated brain aging” (Elastic Net brain age gap: −0.05, CI: − 0.06 - −0.03; XGBoost brain age gap: −0.05, CI: −0.06 - −0.03). However, the volumetric 9 analyses suggested that this was due to the brain age models misclassifying the substantial level effects (hippocampal volume: 0.13, CI: 0.11 - 0.14; total brain volume: 0.21, CI: 0.20 - 0.23) as aging differences. As expected, the longitudinal analyses (n = 1,922) revealed no significant relationships between birth weight and brain change (hippocampal change: −0.02, CI: −0.08 − 0.04; total brain volume change: −0.01, CI: −0.06 − 0.03).

#### Dissociation 2: Individual differences in slope but not level

Tau levels for each participant were derived from PET tau scans. Since not all PET scans were attained at the same imaging session as the structural MRIs we used Sample Iterative Approximation (SILA) modeling to estimate TAU-PET at MRI scan time [49]. For each MRI session we used a threshold of 1.11 Standardized Uptake Value Ratio (SUVR) to determine tau positivity [50]. A logistic regression model was used to examine the relationship between brain age gap and tau positivity, correcting for age and scanner site (n = 1199 for cross-sectional analyses, n = 856 for longitudinal analyses). The results are presented in Fig. 3**f**. As predicted from the model analysis and the simulation experiment, brain age gap showed relatively low sensitivity to tau positivity (Elastic Net brain age gap OR = 1.21, CI: 1.06 − 1.38; XGBoost brain age gap OR = 1.23, CI: 1.06 − 1.42). This was in contrast to simple volumetric change measures, which showed excellent sensitivity to tau positivity (hippocampal volume change: OR = 2.41, CI: 1.65 - 3.52; total brain volume change: OR = 2.97, CI: 1.51 - 5.85). Even the raw volumes showed numerically higher sensitivity to tau positivity than the brain age models (hippocampal volume: OR = 1.47, CI: 1.28 - 1.70; total brain volume: OR = 1.54, CI: 1.07 - 2.20), a difference which was significant in the case of hippocampus (p = 0.0034 vs. Elastic Net brain age gap). This follows directly from the mechanism described above: brain age combines features using weights that are inversely related to each feature’s total variance, so features with large between-person variance in aging rate, exactly those most informative about individual differences, are systematically down-weighted, whereas a single volumetric measure retains its full slope-related variance.

## 3 Discussion

We show that conventional brain age models by design are poorly suited to detect individual differences in brain change, while conferring a high risk of false positive results by misclassifying stable individual differences as differences in aging rate. Using simulations, we demonstrate that features reflecting stable between-person differences are systematically upweighted in age-prediction models as long as they correlate with chronological age, whereas features that primarily capture between-person differences in longitudinal change are downweighted. This pattern was mirrored in real data: brain age models tended to indicate accelerated aging in a setting where group differences were primarily due to stable level effects, while showing low sensitivity to brain atrophy in a setting where real longitudinal differences dominate. Importantly, this bias will increase with more accurate cross-sectional models - as chronologicalage prediction improves, features carrying information about individual differences in change will be even more strongly down-weighted. This creates a paradox where less accurate brain age models in some cases are more sensitive to individual differences in brain aging, which constitutes a special case of the reliability paradox [51]. Our results strongly suggest that conventional brain age models should not be interpreted as measures of individual differences in brain aging, and we caution against treating brain age gap as a proxy for accelerated or decelerated brain aging without additional evidence. Importantly, these limitations extend beyond aging to the interpretation of brain health: because brain age models to a large extent reflect stable anatomical differences, they can label individuals with lifelong smaller brain volumes as having poorer “brain health,” while remaining relatively insensitive to ongoing neurodegenerative change. We also note that the exact same concerns apply when using brain age models to describe development. We suggest more appropriate approaches can be used to detect individual differences in brain aging from cross-sectional data when longitudinal information is unavailable.

### 3.1 Empirical consequences of the design mismatch

Birth weight was associated with lower brain age gap, which could readily be interpreted as “younger” or more resilient brain aging in individuals with a more favourable prenatal environment [21]. However, when we examined simple volumetric measures, birth weight was primarily related to between-person differences, with little evidence of differences in aging rate, consistent with recent large-scale longitudinal studies [36]. The longitudinal null in our data rules out the alternative interpretation that low birth weight produces a real aging effect through shared developmental-vulnerability mechanisms. Whatever the brain age model is detecting, it is unlikely to be ongoing differences in aging rate, because none are observable longitudinally. This illustrates how brain age can transform level effects established before adulthood into apparent aging effects, creating a substantial risk of false-positive inferences about accelerated or decelerated brain aging.

Schizophrenia provides a particularly important example because the inferential stakes are high. The disorder is widely conceptualized as neurodevelopmental [52], but longstanding theories and empirical work have also raised the possibility of progressive brain changes after onset [53, 54]. Longitudinal studies have indeed reported greater brain change over time in patients than controls in some samples [55], making this a clinically and etiologically important question. In this context, the large ENIGMA schizophrenia study reported a mean brain age difference of 3.55 years between patients and controls and framed this as evidence of “advanced structural brain ageing” [8]. However, the reported brain age gap was not significantly associated with illness duration, age of onset, symptom severity, or antipsychotic treatment/dose. Our results suggest that such a pattern could both be compatible with stable case-control level differences in brain structure, and with mixtures of level and change that cannot be disentangled by cross-sectional brain age alone. The interpretational risk is amplified by the fact that brain age gap in schizophrenia appears to be driven strongly by global gray-matter volume [56], and global morphometric measures are precisely the kinds of features that previous studies [32] and our current results indicate are often dominated by stable between-person differences. This is especially relevant given prior ENIGMA reports of robust case-control morphometric differences in schizophrenia, including widespread cortical [57] and subcortical [58] abnormalities, often interpreted as reflecting neurodevelopmental and other trait-like influences in addition to any ongoing progression. Our point is therefore not that prior brain age findings in schizophrenia are wrong, but that group differences in brain age can easily be overinterpreted as evidence of accelerated brain aging when level-based explanations remain likely. Hence, higher brain age in patients with a schizophrenia diagnosis cannot be interpreted to suggest they experience accelerated aging without additional evidence. This issue is of course not specific to schizophrenia, but is particularly important in cross-sectional studies of conditions with a neurodevelopmental or genetic foundation producing stable differences in brain anatomy. The distinction between level and change directly shapes scientific and clinical inference. If stable anatomical differences are interpreted as accelerated brain aging, etiological models may shift toward progressive-degeneration accounts and underestimate early-life, neurodevelopmental, or genetic contributions [35].

Unfortunately, our theoretical and empirical analyses demonstrate that false positive results about brain aging are not the only problem of brain age models - they also tend to lack sensitivity to detect real and clinically meaningful biological change. Tau PET accumulation, a biomarker mechanistically linked to neurodegeneration [59], was associated with brain decline, yet brain age gap showed low sensitivity to this tau-related change. By contrast, longitudinal change in hippocampal and total brain volume showed consistent associations, in line with prior work [39]. Previous studies have reported associations between brain age measures and Alzheimer’s disease biomarkers, including tau (e.g., [60]), but we find that even a single, cross-sectional hippocampal volumetric measure was significantly more sensitive to tau-related neurodegeneration than the brain age models. This result follows from the suppressive weighting derived above: brain age penalizes features with high slope-related variance, whereas a single volume does not. Our modeling analysis hence shows that this relative insensitivity is inherent in the construction of the brain age models, and should increase further as age-prediction models become more accurate. This is exactly what was observed in a recent study finding higher sensitivity for disease detection in less accurate brain age models [61]. Although the present analyses focus on aging, the same conceptual problems apply to brain age models in development, and with greater stakes, since developmental trajectories show substantial between-individual variation in timing and rate, and detecting precisely that variation (e.g., developmental delay or accelerated maturation) is a primary use of such models. The training objective penalizes the very signal these applications are intended to recover.

### 3.2 Suggestions for ways forward

In addition to the problems discussed above, collapsing heterogeneous aging patterns into a single brain age estimate entails substantial information loss, obscuring potentially important insights [62–64]. Longstanding evidence demonstrates that brain aging reflects different influences and is biologically heterogeneous across regions, systems, and modalities [36, 65–70], and compressing this variation into a single global age score easily reduces interpretability and dilutes signals from mechanistically distinct aging processes. This distinction remains critical even when the aim is group comparison rather than mechanistic explanation. As we show, there is an inherent tension between models optimized to explain shared age-related variance (i.e., chronological age prediction) and models intended to detect individual differences in aging-related change [51]. Hence, brain age models function primarily as a relabeled composite of anatomical variation, rather than a measure of aging dynamics, which is problematic due to the semantic association between “brain age” and “brain aging”. As highlighted above, renaming brain age gap as indexing “brain health” does not alleviate the concerns we raise since the insensitivity to individual variation still remains. Given the conceptual and clinical consequences of conflating offset with change, the next question is not whether cross-sectional data can inform brain aging, but how they should be used appropriately. Here the central issue is not how well a model fit [61, 71, 72], but what the model is optimized to predict, and we argue that brain age models are optimized for the wrong target. This reframing points directly to a practical question: what should such models be optimized to predict instead?

With conventional structural MRI measures and study designs, longitudinal estimates are often limited by low reliability, especially in short follow-up studies [73]. In fact, for some features and age ranges, the amount of change-related information recoverable from short-term longitudinal data may be so limited that well-chosen cross-sectional measures, especially those with a higher proportion of accumulated change signal, can carry as much or more information about aging-related variation [32]. We suggest that progress will require cross-sectional models that are explicitly designed to capture aging-relevant variation, do not capture lifelong differences and are validated against meaningful biological change. Third-generation biological aging clocks follow this principle by predicting downstream indicators of aging-related decline rather than chronological age [74–76].

An analogous strategy in neuroimaging would be to train models that predict longitudinal change from baseline brain features [74]. For example, canonical correlation analysis could be used to relate multivariate baseline brain measures to subsequent changes in those same features. The resulting weight vector would define a cross-sectional summary score that maximally predicts future change. Because individual differences in longitudinal change reflect differences in aging rates over the observation interval, such a score can be interpreted as a cross-sectional proxy for accelerated aging. Relatedly, as individual aging rates imply that the variation around the mean should expand with age (“delta expansion”), this can be exploited when longitudinal data is limited. By considering a set with features that show delta expansion, one can obtain a combined measure of individual-age sensitive measures without longitudinal data [33]. This approach differs fundamentally from conventional brain age prediction: instead of maximizing accuracy for chronological age, the model is optimized to capture inter-individual differences in aging-related change.

While variability in many brain-derived features is likely dominated by stable between-subject differences rather than differences in aging rate in young and middleaged [32], in later life, age-related change contributes a larger proportion of the overall variance. In these settings, age-normed deviations of selected features could provide some information about accelerated aging. For example, lateral ventricular volume is sensitive to ongoing atrophy and may serve as a proxy marker related to accelerated aging in older individuals. Similarly, when strong regional hypotheses exist, such as hippocampal atrophy in Alzheimer’s disease, normed deviations in the affected region may to some extent provide interpretable and clinically relevant markers. More global measures may also be informative: for example, total brain volume adjusted for intracranial volume has been suggested as an indicator of cumulative lifetime atrophy [77]. Such feature-wise normative modeling may provide a more transparent alternative to brain age models, particularly in older populations or disease contexts where ongoing structural change is substantial.

## 4 Conclusion

While brain age has an immediate appeal as an intuitive metric of brain aging, summarizing complex data in a single number, our analyses show that conventional brain age models cannot deliver on this promise, because the objective they optimize is fundamentally misaligned with the quantity they are taken to measure. We argue the field should reorient towards a target aligned with individual differences in brain change - and validated against longitudinal data where possible - by predicting future change directly, by exploiting the expansion of inter-individual variance that real aging produces, or by using region-specific normative deviations where ongoing atrophy dominates the signal. The path forward is not to abandon cross-sectional imaging as a window on aging, but to ask it the right question.

## Data Availability

The raw data were gathered from many different datasets. Different agreements are required for each dataset. Most datasets are openly available with prespecified data usage agreements. For some datasets, such as UKB, fees may apply. See the Supplementary Material for a detailed description of how to access the data.

## 5 Acknowledgements

This work was supported by South-Eastern Norway Regional Health Authority (to *EG* [HSØ-2021079]), the Department of Psychology, University of Oslo (to *KW, AF*), the Norwegian Research Council (to *KW, AM, DP* [ES694407]) and the project has received funding from the European Research Council’s Starting Grant scheme (to *AF* [283634, 725025] and *KW* [313440]).

## 6 Code and Data Availability

The raw data were gathered from many different datasets. Different agreements are required for each dataset. Most datasets are openly available with prespecified data usage agreements. For some datasets, such as UKB, fees may apply. See the Supplementary Material for a detailed description of how to access the data. The code is available at https://github.com/EdvardGrodem/Reconsidering-Brain-Age.

## 7 Author contributions

EOSG contributed to the theoretical analysis, experimental validation, and writing. SMS contributed to the theoretical analysis and writing. DV-P contributed to the experimental validation and writing. MLE contributed to the experimental validation and writing. ADNI contributed data. KBW contributed to funding acquisition and writing. AMF contributed to funding acquisition, experimental validation, and writing.

## A Appendix

### A.1 Theoretical Study of Brain Age

Here we describe in detail why brain age models suppress individual longitudinal change. Imagine that we follow a cohort throughout life. At the initial age of *t* = *t*_0_ (which could be any age, say 18 years) of our cohort, the features ***X*** in our cohort have a mean value ***µ***(*t*_0_) and a covariance matrix **Σ**(*t*_0_). As the cohort ages, the mean will change smoothly, and so will the covariance matrix. This means that these are continuous functions of age: ***µ***(*t*) and **Σ**(*t*) (see Fig. 4). The covariance matrix will at any time be the sum of three matrices, **Σ**(*t*) = ***R*** + **Σ**_0_(*t*) + **Σ**_*r*_(*t*). ***R*** is the variance that is not-persistent over the timespan over which we are interested in measuring changes. This could be attributed to hourly variations due to activity, hydration, time-of-day, scanner noise, and variability from the processing software. Here we assume that it is independent of age, to make it easier to separate from the two other sources of variance. It is possible that ***R*** is age-dependent, but this would not change our conclusions. **Σ**_0_(*t*) is the covariance in the data that stems from the baseline effect at *t*_0_. The various elements of **Σ**_0_(*t*) may change with age and represent the dynamics that can be predicted from the initial age. **Σ**_0_(*t*) represents the level effects. **Σ**_*r*_(*t*) is the covariance that originates from random or non-predictable influences since *t*_0_. For instance, this may be caused by disease, lifestyle, genetics, accidents and much more. If this alteration from the norm is in the same direction as the influence of aging, we can interpret this change as accelerated aging. Thus **Σ**_*r*_(*t*) captures the slope effects.

**Fig. 4.**
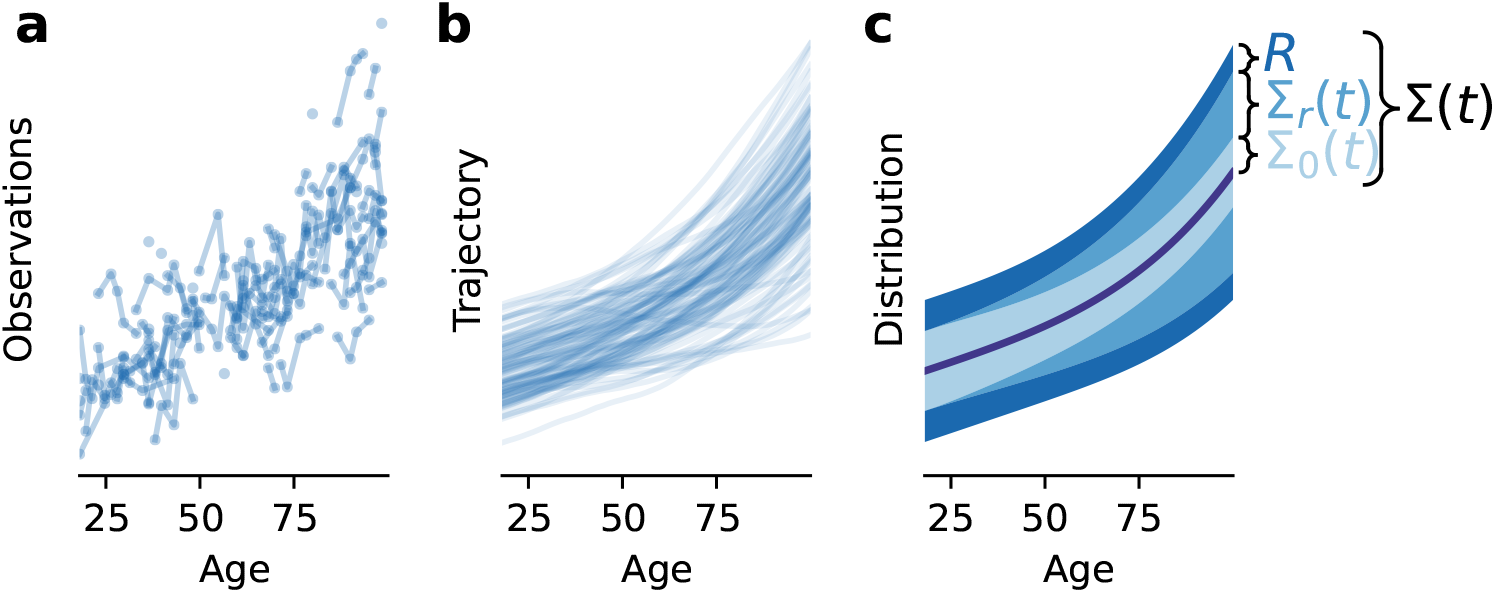
Decomposition of the variance of a dynamical system. The partial observation **a** of the true trajectories **b**. Given enough data we can estimate the true age dependent distribution shown in **c**. At each age the variance around the mean is composed of three sources of variance: the measurement noise ***R***, the variance due to variation in initial condition **Σ**_0_, and the variance from different aging dynamics **Σ**_*r*_.

Most brain age models are created by letting a model *m*(***x***) minimize the mean-squared error of chronological age, *t*, to the model’s output age *m*(***x***), which is termed the “brain age”.

Commonly, a linear model is used. Here we ignore regularizing terms used in models such as Lasso and ridge regression, since these terms only force the coefficients of the model towards zero. The coefficients ***β*** of a linear model are simply given by

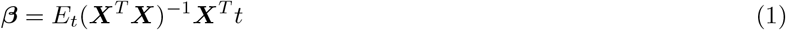

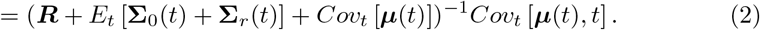

Thus, the marginal contribution of a single feature *x*_*i*_ to the variance in the brain age model at a chronological age *t* is

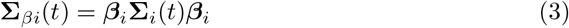

The magnitude of the coefficient of each feature is a weighting of how the mean of the feature varies with age compared to the variance of the feature. This immediately reveals the issue with brain age models. Features with a high amount of accelerated aging, i.e., high expected **Σ**_*r*_(*t*), are downweighted compared to features with very little variance due to accelerated aging **Σ**_*r*_(*t*) *≈* **0**.

We can also investigate the properties of a non-linear model *m*_*nl*_(***x***). We will assume that the model is optimal in the sense that it produces *E* [*t*|***x***] and that it is unbiased such that *E* [*m*_*nl*_(***x*** | *t*)] := *t*, meaning if we take all the participants with age *t* and run our non-linear model on them, the mean brain age will be *t*. This is often a desired property for brain age models, but must be corrected for since *E* [*t* | ***x***] is biased by the age distribution. Note that we are not assuming that the model can perfectly fit the age of the participant, only that it does the best job possible given the brain age objective and the data.

Brain age models are usually very good at predicting the age given the data, with some models reporting down to *~*2.5 year mean absolute error, and the brain age gap will thus be of the same magnitude.

What features is the optimal nonlinear model using when analyzing the brain of an individual at a chronological age of 70, and for instance, predicts the brain to be 68 years or 72 years? To investigate this, we can linearize the model around the mean structure at age 70 (or any other age). The first order approximation of this model around an age *t*_*s*_ is given by

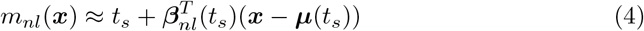

and the brain age gap will be

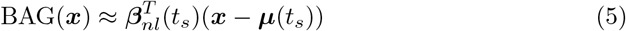

Given that the brain is changing in a smooth fashion, the relative magnitude of coefficients ***β***_*nl*_ tell us the contribution of each feature used by the model. The linearized coefficient is given by

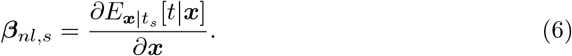

Which we can show is

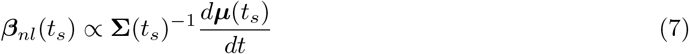

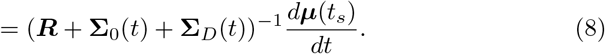

A very similar result to the linear case; however, there are some important distinctions. In the optimal model case, the total variance is only estimated for each age, further amplifying the suppression of features sensitive to individual aging. This is exemplified by in Fig. 2 where the large spread of the F2 makes the optimal model suppress the feature completely at the higher ages, even though this feature has high amounts of signal from individualized accelerated aging.

### A.2 Simulated dynamical-system experiment illustrating suppression of individualized aging effects

To illustrate the theoretical results above, we constructed a simple dynamical-system simulation in which two features evolve across the lifespan with distinct sources of variability. The goal of this experiment is to demonstrate how brain age models preferentially weight features dominated by predictable cohort-level change while suppressing features that primarily reflect individualized longitudinal dynamics.

We simulated a cohort evolving continuously from age 18 to 90 years using a linear Gaussian state-space model. Each individual is described by a four-dimensional latent state vector

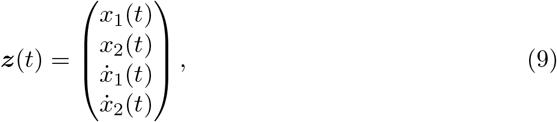

where *x*_1_(*t*) and *x*_2_(*t*) are observable features and 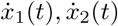 represent their rates of change. The system evolves according to the discrete-time linear dynamics

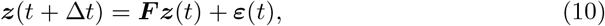

with transition matrix

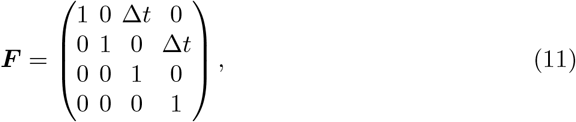

and Gaussian process noise

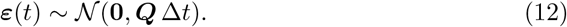

The process noise covariance was chosen such that only the derivative of the second feature contained stochastic variation,

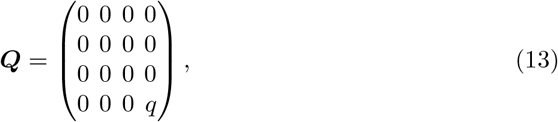

so that feature *x*_2_ accumulates unpredictable subject-specific variation over time, whereas feature *x*_1_ evolves deterministically apart from baseline variation. This construction directly separates predictable cohort-level effects from individualized slope effects in the sense of the decomposition

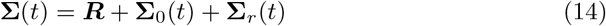

introduced above.

Initial states at age *t*_0_ = 18 were sampled from

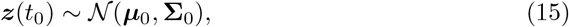

with nonzero baseline variance only in the feature levels,

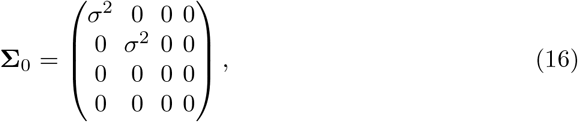

and the same mean inital conditions of

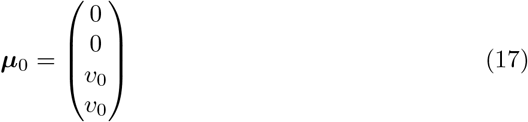

Thus both features start with the same cross-sectional variability and age relation at baseline, but only feature *x*_2_ accumulates additional variance through individualized longitudinal dynamics.

Under this model the cohort mean evolves smoothly with age,

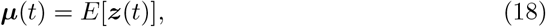

while the covariance evolves according to

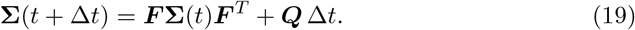

As a consequence, the variance of feature *x*_2_ increases progressively with age due to accumulated process noise, whereas the variance of feature *x*_1_ remains determined by baseline differences between individuals. In terms of the decomposition introduced earlier, feature *x*_1_ is dominated by **Σ**_0_(*t*), while feature *x*_2_ increasingly reflects **Σ**_*r*_(*t*).

To analyse how brain age models respond to these different sources of variation, we computed the locally optimal regression weights implied by the posterior mean estimator

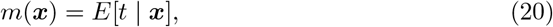

using the expression derived in Proposition 1,

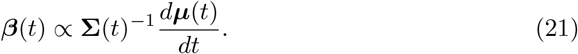

This provides the theoretically optimal feature weights at each age under a Gaussian observation model.

For comparison, we also estimated a global linear brain age model using the time-averaged covariance matrix,

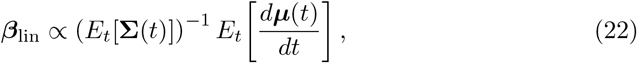

and trained a nonlinear brain age predictor using gradient-boosted decision trees (XGBoost) on synthetic cross-sectional samples drawn from

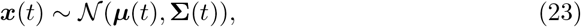

with 100 samples per chronological age. Feature contributions in the XGBoost model were evaluated locally by regressing predicted age on features within each age stratum and computing the proportion of the variance contributed to each feature.

### A.3 Local linear form of the posterior mean under a Gaussian observation model

Here a more formal description is given for a Gaussian continuous observation model.

#### Proposition 1.

***Local approximation of*** *m*(***x***) = 𝔼 [*t*| ***x***]

*Let*

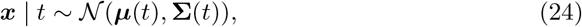

*where* ***µ***(*t*) *and* **Σ**(*t*) *are smooth in t* ∈ ℝ, *and let t have density p*(*t*). *Fix t*_*s*_ *and define*

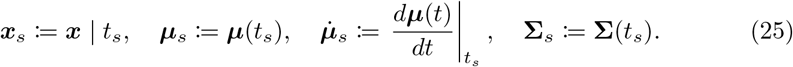

*Let the regression function be*

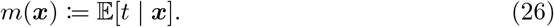

*Assume that around t*_*s*_ *the model is locally unbiased*,

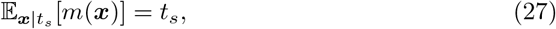

*and assume that p*(*t* | ***x***_*s*_) *is sufficiently concentrated such that within the high-probability region of p*(*t* | ***x***_*s*_), *we may approximate* ***µ***(*t*) *as affine*, **Σ**(*t*) *as constant, and p*(*t*) *as flat, i*.*e*.

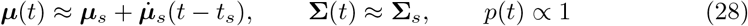

*Then m*(***x***) *admits the local approximation around t*_*s*_ *given by*

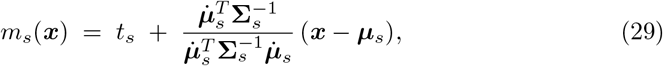

*and the variance of m*(***x***) *under* ***x*** | *t*_*s*_ *is*

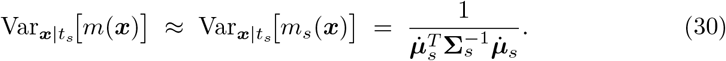

*Proof* Under the stated concentration/linearity assumptions, for ***x*** in the high-posterior region around (*t*_*s*_, ***x***_*s*_),

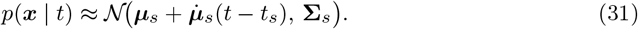

Moreover, since *p*(*t*) is locally flat around *t*_*s*_, we take log *p*(*t*) to contribute (approximately) no *t*-dependence in the local posterior, so

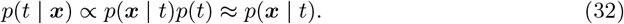

Let *τ* := *t* − *t*_*s*_ and write ***δ*** := ***x*** − ***µ***_*s*_. Then, up to a *τ* -independent normalizing constant,

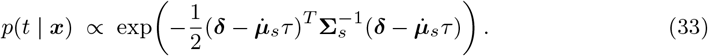

Expand the quadratic form:

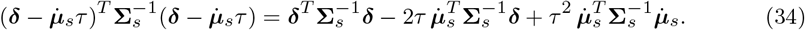

Define

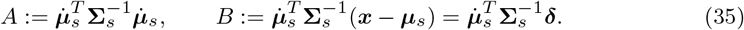

Then, ignoring constants independent of *τ*,

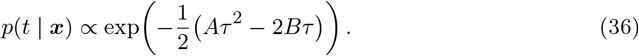

Complete the square:

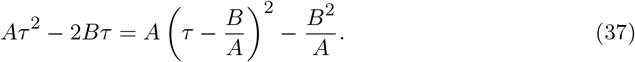

Again we can ignore terms that are independent of *τ* which gives:

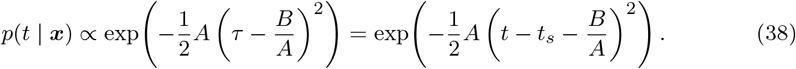

Hence, the posterior *p*(*t*|*x*) is approximately Gaussian with mean *t*_*s*_ + *B/A* and variance 1*/A*,

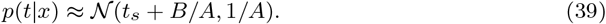

Therefore

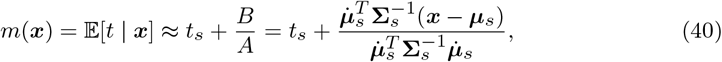

which is (29).

For the variance under ***x*** | *t*_*s*_, use the linear form

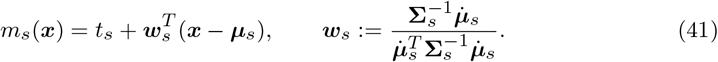

Since ***x*** | *t*_*s*_ *~* 𝒩 (***µ***_*s*_, **Σ**_*s*_),

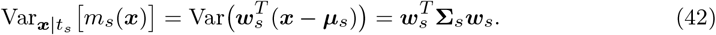

Substituting ***w***_*s*_ gives

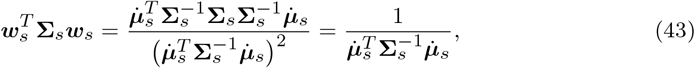

which is (30). □

## References

[1] Cole, J. H. & Franke, K. Predicting age using neuroimaging: innovative brain ageing biomarkers. Trends in Neurosciences 40, 681–690 (2017).

[2] Cole, J. H. et al. Predicting brain age with deep learning from raw imaging data results in a reliable and heritable biomarker 163, 115–124 (2017).

[3] Franke, K. et al. In vivo biomarkers of structural and functional brain development and aging in humans 117, 142–164 (2020).

[4] Legaz, A. et al. The exposome of brain aging across 34 countries. Nature Medicine 1–14 (2026).

[5] Koutsouleris, N. et al. Accelerated brain aging in schizophrenia and beyond: a neuroanatomical marker of psychiatric disorders 40, 1140–1153 (2014).

[6] Nenadic, I., Dietzek, M., Langbein, K., Sauer, H. & Gaser, C. BrainAGE score indicates accelerated brain aging in schizophrenia, but not bipolar disorder 266, 86–89 (2017).

[7] Hajek, T. et al. Brain age in early stages of bipolar disorders or schizophrenia 45, 190–198 (2019).

[8] Constantinides, C. et al. Brain ageing in schizophrenia: evidence from 26 international cohorts via the ENIGMA Schizophrenia consortium 28, 1201–1209 (2023).

[9] Han, L. K. M. et al. Brain aging in major depressive disorder: results from the enigma major depressive disorder working group 26, 5124–5139 (2021).

[10] Kaufmann, T. et al. Common brain disorders are associated with heritable patterns of apparent aging of the brain 22, 1617–1623 (2019).

[11] Lowe, L. C., Gaser, C., Franke, K. & Alzheimer’s Disease Neuroimaging, I. The effect of the apoe genotype on individual BrainAGE in normal aging, mild cognitive impairment, and Alzheimer’s disease 11, e0157514 (2016).

[12] Leonardsen, E. H. et al. Genetic architecture of brain age and its causal relations with brain and mental disorders 28, 3111–3120 (2023).

[13] Wen, J. et al. The genetic architecture of multimodal human brain age 15, 2604 (2024).

[14] Jawinski, P. et al. Genome-wide analysis of brain age identifies 59 associated loci and unveils relationships with mental and physical health 5(2025).

[15] Oh, H. S.-H. et al. Plasma proteomics links brain and immune system aging with healthspan and longevity. Nature Medicine 31, 2703–2711 (2025).

[16] Franke, K., Gaser, C., Manor, B. & Novak, V. Advanced BrainAGE in older adults with type 2 diabetes mellitus 5, 90 (2013).

[17] Steffener, J. et al. Differences between chronological and brain age are related to education and self-reported physical activity 40, 138–144 (2016).

[18] Cole, J. H. Multimodality neuroimaging brain-age in UK Biobank: relationship to biomedical, lifestyle, and cognitive factors 92, 34–42 (2020).

[19] de Lange et al. Multimodal brain-age prediction and cardiovascular risk: The Whitehall II MRI sub-study 222, 117292 (2020).

[20] Beck, D. et al. Cardiometabolic risk factors associated with brain age and accelerate brain ageing 43, 700–720 (2022).

[21] Franke, K., Gaser, C., Roseboom, T. J., Schwab, M. & de Rooij, S. R. Premature brain aging in humans exposed to maternal nutrient restriction during early gestation 173, 460–471 (2018).

[22] Luders, E. et al. Potential brain age reversal after pregnancy: Younger brains at 4-6 weeks postpartum 386, 309–314 (2018).

[23] de Lange et al. Population-based neuroimaging reveals traces of childbirth in the maternal brain 116, 22341–22346 (2019).

[24] Moguilner, S. et al. Brain clocks capture diversity and disparities in aging and dementia across geographically diverse populations. Nature Medicine 30, 3646–3657 (2024).

[25] Cole, J. H. et al. Brain age predicts mortality 23, 1385–1392 (2018).

[26] Franke, K., Luders, E., May, A., Wilke, M. & Gaser, C. Brain maturation: predicting individual BrainAGE in children and adolescents using structural MRI 63, 1305–1312 (2012).

[27] Whitmore, L. & Beck, D. Current challenges and future directions for brain age prediction in children and adolescents 16, 7771 (2025).

[28] Wittens, M. M. et al. Brain age as a biomarker for pathological versus healthy ageing–a REMEMBER study. Alzheimer’s Research & Therapy 16, 128 (2024).

[29] Vidal-Pineiro, D. et al. Individual variations in ‘brain age’ relate to early-life factors more than to longitudinal brain change 10(2021).

[30] Dörfel, R. P. et al. Prediction of brain age using structural magnetic resonance imaging: a comparison of clinical utility of publicly available software packages. eBioMedicine 123, 106094 (2026).

[31] Korbmacher, M. et al. Cross-sectional brain age assessments are limited in predicting future brain change 46, e70203 (2025).

[32] Grødem, E. O. et al. Stable individual differences dominate adult brain volume variation until later life. Imaging Neuroscience (2026).

[33] Smith, S. M., Miller, K. L. & Nichols, T. E. Characterising ongoing brain aging and baseline effects from cross-sectional data 3(2025).

[34] Walhovd, K. B. et al. Neurodevelopmental origins of lifespan changes in brain and cognition 113, 9357–9362 (2016).

[35] Walhovd, K. B., Lövden, M. & Fjell, A. M. Timing of lifespan influences on brain and cognition 27, 901–915 (2023).

[36] Walhovd, K. B. et al. Fetal influence on the human brain through the lifespan 12 (2024).

[37] Ingram, D. K. Key questions in developing biomarkers of aging. Experimental Gerontology 23, 429–434 (1988).

[38] Sluiskes, M. H. et al. Clarifying the biological and statistical assumptions of cross-sectional biological age predictors: an elaborate illustration using synthetic and real data. BMC Medical Research Methodology 24, 58 (2024).

[39] La Joie, R. et al. Prospective longitudinal atrophy in Alzheimer’s disease correlates with the intensity and topography of baseline tau-PET. Science Translational Medicine 12, eaau5732 (2020).

[40] Leonardsen, E. H. et al. Deep neural networks learn general and clinically relevant representations of the ageing brain. NeuroImage 256, 119210 (2022).

[41] Chen, T. et al. Xgboost: extreme gradient boosting. R package version 0. 4-2 1, 1–4 (2015).

[42] Mueller, S. G. et al. The Alzheimer’s disease neuroimaging initiative. Neuroimaging Clinics 15, 869–877 (2005).

[43] Park, D. et al. The dallas lifespan brain study: A comprehensive adult lifespan data set of brain and cognitive aging. Scientific Data 12 (2025).

[44] Dagley, A. et al. Harvard aging brain study: dataset and accessibility. NeuroImage 144, 255–258 (2017).

[45] Tremblay-Mercier, J. et al. Open science datasets from PREVENT-AD, a longitudinal cohort of pre-symptomatic Alzheimer’s disease. NeuroImage: Clinical 31, 102733 (2021).

[46] Walhovd, K. et al. Neurodevelopmental origins of lifespan changes in brain and cognition. Proceedings of the National Academy of Sciences 113, 9357–9362 (2016).

[47] Miller, K. et al. Multimodal population brain imaging in the UK Biobank prospective epidemiological study. Nature Neuroscience 19, 1523–1536 (2016).

[48] Smith, S. M., Vidaurre, D., Alfaro-Almagro, F., Nichols, T. E. & Miller, K. L. Estimation of brain age delta from brain imaging. Neuroimage 200, 528–539 (2019).

[49] Betthauser, T. J. et al. Multi-method investigation of factors influencing amyloid onset and impairment in three cohorts. Brain 145, 4065–4079 (2022). URL 10.1093/brain/awac213.

[50] Maass, A. et al. Comparison of multiple tau-PET measures as biomarkers in aging and Alzheimer’s disease. NeuroImage 157, 448–463 (2017).

[51] Hedge, C., Powell, G. & Sumner, P. The reliability paradox: Why robust cognitive tasks do not produce reliable individual differences. Behavior Research Methods 50, 1166–1186 (2018).

[52] Solmi, M. et al. Age at onset of mental disorders worldwide: large-scale meta-analysis of 192 epidemiological studies. Molecular Psychiatry 27, 281–295 (2022).

[53] Olabi, B. et al. Are there progressive brain changes in schizophrenia? a meta-analysis of structural magnetic resonance imaging studies. Biological psychiatry 70, 88–96 (2011).

[54] Delisi, L. The concept of progressive brain change in schizophrenia: Implications for understanding schizophrenia. Schizophrenia bulletin 34, 312–21 (2008).

[55] DeLisi, L. E. et al. Schizophrenia as a chronic active brain process: a study of progressive brain structural change subsequent to the onset of schizophrenia. Psychiatry Research: Neuroimaging 74, 129–140 (1997).

[56] Ballester, P. L. et al. Gray matter volume drives the brain age gap in schizophrenia: a SHAP study. Schizophrenia 9, 3 (2023).

[57] van Erp, T. G. et al. Cortical brain abnormalities in 4474 individuals with schizophrenia and 5098 control subjects via the enhancing neuro imaging genetics through meta analysis (ENIGMA) consortium. Biological Psychiatry 84, 644–654 (2018).

[58] van Erp, T. G. et al. Subcortical brain volume abnormalities in 2028 individuals with schizophrenia and 2540 healthy controls via the ENIGMA consortium. Molecular Psychiatry 21, 547–553 (2016).

[59] Ballatore, C., Lee, V. M.-Y. & Trojanowski, J. Q. Tau-mediated neurodegeneration in alzheimer’s disease and related disorders. Nature reviews neuroscience 8, 663–672 (2007).

[60] Cumplido-Mayoral, I. et al. Biological brain age prediction using machine learning on structural neuroimaging data: Multi-cohort validation against biomarkers of Alzheimer’s disease and neurodegeneration stratified by sex. eLife 12, e81067 (2023).

[61] Schulz, M.-A., Siegel, N. T. & Ritter, K. Brain-age models with lower age prediction accuracy have higher sensitivity for disease detection. PLOS Biology 23, 1–14 (2025). URL 10.1371/journal.pbio.3003451.

[62] Smith, S. M. et al. Brain aging comprises many modes of structural and functional change with distinct genetic and biophysical associations 9 (2020).

[63] Cox, S. R. et al. Three major dimensions of human brain cortical ageing in relation to cognitive decline across the eighth decade of life. Molecular Psychiatry 26, 2651–2662 (2021).

[64] Chen, C.-H. et al. Hierarchical genetic organization of human cortical surface area. Science 335, 1634–1636 (2012).

[65] Raz, N. et al. Regional brain changes in aging healthy adults: general trends, individual differences and modifiers 15, 1676–1689 (2005).

[66] Shaw, P. et al. Neurodevelopmental trajectories of the human cerebral cortex. Journal of Neuroscience 28, 3586–3594 (2008).

[67] Fjell, A. M. et al. Development and aging of cortical thickness correspond to genetic organization patterns 112, 15462–15467 (2015).

[68] Nyberg, L. et al. Educational attainment does not influence brain aging 118 (2021).

[69] Fjell, A. M. et al. The genetic organization of longitudinal subcortical volumetric change is stable throughout the lifespan. eLife 10, e66466 (2021).

[70] Fjell, A. M. et al. Reevaluating the role of education on cognitive decline and brain aging in longitudinal cohorts across 33 Western countries. Nature Medicine 31, 2967–2976 (2025).

[71] Bashyam, V. M. et al. MRI signatures of brain age and disease over the lifespan based on a deep brain network and 14 468 individuals worldwide. Brain 143, 2312–2324 (2020).

[72] Hahn, T. et al. From ‘loose fitting’ to high-performance, uncertainty-aware brainage modelling. Brain 144, e31–e31 (2021).

[73] Vidal-Pineiro, D. et al. Vulnerability to memory decline in aging revealed by a mega-analysis of structural brain change 16, 11488 (2025).

[74] Whitman, E. T. et al. Dunedinpacni estimates the longitudinal pace of aging from a single brain image to track health and disease. Nature Aging 5, 1619–1636 (2025).

[75] Raffington, L. & Belsky, D. W. Integrating DNA methylation measures of biological aging into social determinants of health research. Current Environmental Health Reports 9, 196–210 (2022).

[76] Belsky, D. W. et al. DunedinPACE, a DNA methylation biomarker of the pace of aging. eLife 11, e73420 (2022).

[77] Fürtjes, A. E. et al. Measurement characteristics and genome-wide correlates of lifetime brain atrophy estimated from a single MRI. Nature Communications 16, 6725 (2025).

