## Supplementary Information for "Reconsidering Brain Age: Why Age-Prediction Models Fail as Measures of Brain Aging"

### 1 Data sources

#### 1.1 ADNI

The Alzheimer's Disease Neuroimaging Initiative (ADNI) [1] is a multi-site project led by Doctor Michael W. Weiner to assess the progression of mild cognitive impairment (MCI) and early Alzheimer's Disease (AD), combining imaging, clinical and other biological markers, and neuropsychological and clinical assessments over time. For more information, visit https://adni.loni.usc.edu/about/. The age range for the participants is 55-90 years. In-detailed general inclusion and exclusion criteria are described elsewhere [2]. All participants signed an informed consent form and the protocols were approved by the corresponding regional ethical committees in the US and Canada. The present study includes participants from ADNI 1, ADNIGO, ADNI2, and ADNI 3 with at least one tau PET scan.

#### 1.2 PreventAD

The Pre-symptomatic Evaluation of Experimental or Novel Treatments for AD (PreventAD) [3] is a retrospective, long-term study that follows cognitively healthy older individuals with a familiar history of AD. It includes participants enrolled either from an observational cohort or the clinical trial of PreventAD. This study comprises MRI images, blood and CSF samples, and clinical and neuropsychological assessments. Participants in the study had to be at least 60 years old, had $\geq 6$ years of education, and they needed to be cognitively unimpaired at baseline. The Montreal Cognitive Assessment (MoCA) and CDR scales were used to assess cognitive abilities, and participants were considered cognitively intact if their MoCA scores were $\geq 26/30$ or their CDR was $= 0$. Other exclusion criteria at baseline included medical conditions that prevented longitudinal participation or medical contraindications to MRI, use of acetylcholinesterase inhibitors, other approved prescription cognitive enhancers, hypertension, or substance abuse. The inclusion and exclusion criteria have been previously described in detail [3]. The protocols, consent forms, and study procedures were approved by the McGill Institutional Review Board and the Douglas Mental Health University Institute Research Ethics Board. Observations with RBANS $>$ 1SD below the mean and probable MCI, as evaluated by a clinician, were excluded. The present study used data from all subjects with at least one tau PET scan.

#### 1.3 DLBS

The Dallas Lifespan Brain Study (DLBS) [4] is a longitudinal, multi-modal neuroimaging study of the aging mind initiated in 2008. Participants returned for two additional waves of data collection, with an approximate interval of 4–5 years between waves. The DLBS includes structural MRI, diffusion MRI, functional MRI, amyloid PET, tau PET, comprehensive cognitive assessments, and psychosocial measures. The study was designed to investigate brain aging, Alzheimer's disease-related PET biomarkers, and cognition across the adult lifespan, with participants ranging from 20–90 years of age. It was also among the earlier cognitive aging studies to include in vivo measurement of tauopathy using PET imaging with the radiotracer AV-1451, also known as flortaucipir.

Participants were recruited in two cohorts with partly different inclusion and exclusion criteria. In the first cohort, participants were required to be right-handed, fluent in English, between 20 and 89 years old at Wave 1, have at least a 10th grade education, and have a Mini-Mental State Examination (MMSE) score $\geq 26$. Exclusion criteria included major psychiatric or neurological disorders, recent chemotherapy, coronary bypass, history of substance abuse, central nervous system disease or brain injury, immune, kidney, liver, or pulmonary disorders, corrected vision poorer than 20/30, insulin-dependent diabetes, use of sedatives, benzodiazepines, or anti-psychotics, high caffeine consumption, high cholesterol, blood pressure above 160/90, BMI $\geq 35$, recent recreational drug use, or contraindications to MRI. The second cohort included participants aged 50–89 years at Wave 1 who were enrolled in the Amyloid PET Scan Study, were right-handed, fluent in English, had at least a 9th grade education, and met additional education and health-related criteria. At Waves 2 and 3, participants were required to complete testing within a re-test interval of 2.5–11 years, and the MMSE exclusion cutoff was lowered to $< 22$ to allow potential cognitive decline to be captured.

Tau PET imaging at Waves 2 and 3 used $^{18}$F-AV-1451, also known as flortaucipir.

#### 1.4 HABS

The Harvard Aging Brain Study (HABS) [5] is an ongoing, long-term observational study that aims to enhance our understanding of brain aging and the early stages of Alzheimer's disease. The study collects PET, MRI data, neuropsychological and clinical assessments. The age range was between 50 and 90 years at the time of baseline assessment and all patients were considered non-clinically impaired at the start of the study. Further participants had a CDR score of 0, MMSE score $\geq 25$, $< 11$ on the Geriatric Depression Scale, and scores above age- and education-adjusted cutoffs on the 30-Minute Delayed Recall of the Logical Memory Story A to be included in the study. Participants with a history of alcoholism, drug abuse, head trauma, or current serious medical/psychiatric illness were excluded. Further details can be found elsewhere [5]. All participants signed an informed consent form and the protocol was approved by the Partners Healthcare Human Research Committee. The present study used data from all subjects with at least one tau PET scan. We used data from HABS data release 2.20, retrieved in August 2022 via https://habs.mgh.harvard.edu/.

#### 1.5 UKB

The UK Biobank (UKB) (https://www.ukbiobank.ac.uk/about-biobank-uk/) is a major national and international health resource with the aim of improving the prevention, diagnosis and treatment of a wide range of illnesses. UK Biobank recruited ≈500,000 people aged between 40-69 years in 2006-2010 from across the country to take part in this project (Guggenheim et al., 2015). Potential participants were identified through National Health Service (NHS) registers according to being aged 40-69 and living within a reasonable traveling distance of an assessment center. Assessment centers are located in accessible and convenient locations with a large surrounding population. Participants have undergone measures and provided samples and detailed information about themselves and agreed to have their health followed. The study sample was drawn from the UK Biobank neuroimaging branch [6] and conducted under data application number 32048. Only individuals with longitudinal MRI data were used in this study. The subsample used in this study consists of the participants in the first wave of longitudinal imaging. Participants signed an informed consent and the protocols were approved by the North West Multi-Center Research Ethics Committee [MREC]; see also https://www.ukbiobank.ac.uk/the-ethics-and-governance-council.

#### 1.6 LCBC

The Center for Lifespan Changes in Brain and Cognition cohort (LCBC, Oslo) [7] consists of cognitively healthy, community-dwelling participants across the lifespan and is drawn from studies coordinated by the LCBC Research Group (LCBC www.oslobrains.no), approved by a Norwegian Regional Committee for Medical and Health Research Ethics. Written informed consent was obtained from all participants. The samples were recruited by a variety of methods such as newspapers and webpage ads. Most participants were recruited for observational studies, some currently ongoing, while a minority were recruited to enter into cognitive training. Written informed consent was obtained from all adult participants. All participants had to undergo a standardized health interview before being included in the study, and those with a history of neurological or psychiatric conditions or who reported concerns about their cognitive function were excluded. Additionally, all participants over the age of 40 years were required to score at least 25 on the Mini-Mental State Examination. The LCBC cohort was part of the Lifebrain obtained as part of the Lifebrain consortium [8].

**Table 1**: MRI-derived features used in the brain age models. Corresponding right- and left-hemisphere features were combined by summation prior to modelling.

| Region / structure | Area | Thickness | Volume |
|---|---|---|---|
| bankssts | ✓ | ✓ | ✓ |
| caudalanteriorcingulate | ✓ | ✓ | ✓ |
| caudalmiddlefrontal | ✓ | ✓ | ✓ |
| cuneus | ✓ | ✓ | ✓ |
| entorhinal | ✓ | ✓ | ✓ |
| frontalpole | ✓ | ✓ | ✓ |
| fusiform | ✓ | ✓ | ✓ |
| inferiorparietal | ✓ | ✓ | ✓ |
| inferiortemporal | ✓ | ✓ | ✓ |
| insula | ✓ | ✓ | ✓ |
| isthmuscingulate | ✓ | ✓ | ✓ |
| lateraloccipital | ✓ | ✓ | ✓ |
| lateralorbitofrontal | ✓ | ✓ | ✓ |
| lingual | ✓ | ✓ | ✓ |
| medialorbitofrontal | ✓ | ✓ | ✓ |
| middletemporal | ✓ | ✓ | ✓ |
| paracentral | ✓ | ✓ | ✓ |
| parahippocampal | ✓ | ✓ | ✓ |
| parsopercularis | ✓ | ✓ | ✓ |
| parsorbitalis | ✓ | ✓ | ✓ |
| parstriangularis | ✓ | ✓ | ✓ |
| pericalcarine | ✓ | ✓ | ✓ |
| postcentral | ✓ | ✓ | ✓ |
| posteriorcingulate | ✓ | ✓ | ✓ |
| precentral | ✓ | ✓ | ✓ |
| precuneus | ✓ | ✓ | ✓ |
| rostralanteriorcingulate | ✓ | ✓ | ✓ |
| rostralmiddlefrontal | ✓ | ✓ | ✓ |
| superiorfrontal | ✓ | ✓ | ✓ |
| superiorparietal | ✓ | ✓ | ✓ |
| superiortemporal | ✓ | ✓ | ✓ |
| supramarginal | ✓ | ✓ | ✓ |
| transversetemporal | ✓ | ✓ | ✓ |
| Accumbens-area | ✓ | | |
| Lateral-Ventricle | | | ✓ |
| Inf-Lat-Vent | | | ✓ |
| Cerebellum-White-Matter | | | ✓ |
| Cerebellum-Cortex | | | ✓ |

*Continued on next page*

| Region / structure | Area | Thickness | Volume |
|---|---|---|---|
| Thalamus-Proper | | | ✓ |
| Caudate | | | ✓ |
| Putamen | | | ✓ |
| Pallidum | | | ✓ |
| 3rd-Ventricle | | | ✓ |
| 4th-Ventricle | | | ✓ |
| Brain-Stem | | | ✓ |
| Hippocampus | | | ✓ |
| Amygdala | | | ✓ |
| CSF | | | ✓ |
| VentralDC | | | ✓ |
| vessel | | | ✓ |
| choroid-plexus | | | ✓ |
| SubCortGrayVol | | | ✓ |
| TotalGrayVol | | | ✓ |
| SupraTentorialVol | | | ✓ |
| EstimatedTotalIntraCranialVol | | | ✓ |

### 2 Method details of empirical demonstration

Here, we provide a more detailed description of the empirical demonstration. This analysis is intended as a proof of concept, showing the consequences of using brain age models to quantify accelerated ageing. It is not intended as a reference analysis of the associations between birth weight or tau levels and brain measures.

#### 2.1 MRI

Structural MRI data were obtained from ADNI, DLBS, HABS, Prevent-AD, LCBC, and UK Biobank (UKB). All MRI data, except UKB data, were processed on a local secure server using the longitudinal stream of FreeSurfer v7.1.0 to extract measures of cortical area, cortical thickness, cortical volume, and subcortical/global volume. The preprocessing of the UKB sample is given in detail in the UKB documentation: https://biobank.ndph.ox.ac.uk/ukb/ukb/docs/brain_mri.pdf. Subjects younger than 50 years and older than 90 years were excluded from the analysis.

To reduce collinearity between hemispheric features, corresponding features from the right and left hemispheres were combined prior to modelling. The complete set of MRI-derived features used in the brain age models is listed in Supplementary Table 1.

We estimated imaging-site effects using a linear model in `statsmodels` with the following formula:

```
feature ~ cr(age, df=5) + C(site) + ICV
```

where `feature` denotes the MRI-derived feature being corrected, `cr(age, df=5)` denotes a cubic regression spline for age with 5 degrees of freedom, `site` denotes

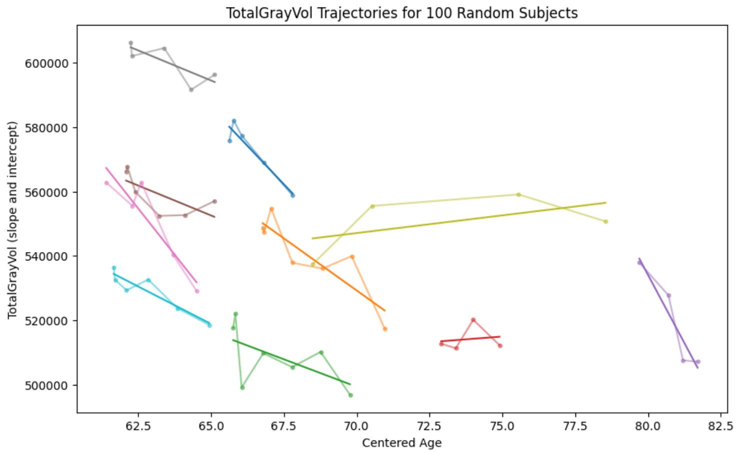


**Fig. 1** Examples of longitudinal grey matter volume trajectories used to estimate subject-specific slopes.

imaging site, and `ICV` denotes intracranial volume. The estimated site effect was then subtracted from each session. This procedure corrected imaging features for site effects while accounting for non-linear age-related variation.

To obtain longitudinal estimates of change, a linear model was fitted separately for each subject using the site-corrected sessions, providing a slope and intercept for each subject. The slope was used in the longitudinal analyses. Examples are shown in Supplementary Fig. 1. The upper and lower 0.1% of the feature values and the upper and lower 1% of the slopes were clipped to reduce the influence of outliers. All features were z-scored after outlier clipping.

For cross-sectional analyses, one session per participant was selected as the session closest to the participant's mean age across visits. This reduces regression-to-the-mean effects when cross-sectional measures are analysed together with longitudinal slope estimates.

#### 2.2 Brain age models

Brain age prediction models were trained on cross-sectional data from UK Biobank. In total, 42,590 UKB participants were used for model training and evaluation. We trained both an Elastic Net regression model and a non-linear XGBoost model to predict chronological age from the MRI-derived features.

The UKB data were divided into five folds. For each split, three folds were used for training, one fold for validation, and one fold for testing. Hyperparameter tuning was performed on the validation set of the first training-validation-test split, and the same hyperparameters were then used for the remaining training-validation-test splits. By rotating which fold was used as the test fold, we obtained out-of-sample brain age estimates for all UKB participants without data leakage. For the non-UKB datasets, an ensemble of the UKB-trained brain age models was used.

For the Elastic Net model, the hyperparameter search was performed over a grid of alpha values ranging from $10^{-8}$ to $10^{-3}$ with 10 steps, and l1-ratio values ranging from 0 to 1 with 20 steps.

For the XGBoost model, the hyperparameter search was performed over the following grid:

```
n_estimators_hyperparams = [100, 300, 600, 1000]
max_depth_hyperparams = [3, 5, 7]
learning_rate_hyperparams = [0.01, 0.1, 0.2]
```

Because brain age predictions typically show systematic age-dependent bias [9], predicted ages were corrected using a Gaussian kernel approach. Specifically, the expected brain-age prediction given chronological age was estimated in the training data as:

$$E[m(\boldsymbol{X}(t)) \mid t] \approx \frac{\sum_i m(\boldsymbol{x}_i) \exp\left(-\frac{1}{2}\left(\frac{t_i - t}{\sigma_k}\right)^2\right)}{\sum_i \exp\left(-\frac{1}{2}\left(\frac{t_i - t}{\sigma_k}\right)^2\right)}, \tag{1}$$

where $m$ is the brain age model, $\boldsymbol{x}_i$ denotes the features from session $i$, $t_i$ is the chronological age at that session, $\sigma_k$ is the kernel spread, and $t$ is the central chronological age. We used a Gaussian kernel with standard deviation $\sigma_k = 2$ years. Brain age predictions were then bias-corrected by subtracting this estimated expected prediction. The brain age gap was computed as the difference between the bias-corrected predicted age and chronological age.

For comparison with simpler structural MRI measures, we selected hippocampal volume, total grey matter volume, and the longitudinal slope of each measure for each participant.

#### 2.3 Birth weight association

We used self-reported birth weight measures from UK Biobank and registry-based birth weight data from LCBC. Outliers were removed by excluding birth weights above 6 kg and below 1 kg.

We tested whether birth weight was associated with brain age gap or volumetric measures. Birth weight is fixed at birth and therefore cannot itself change with age. Thus, an association between birth weight and brain age gap would be interpreted as a test of whether brain age models misclassify stable individual differences in brain structure as differences in brain aging.

To estimate the relationship between birth weight and brain age gap, separate models were fitted for the Elastic Net and XGBoost brain age gaps using the following formula:

```
bw ~ bs(age, df=5) + brain_age_gap + C(site) + C(sex_female)
```

where `brain_age_gap` denotes the brain age gap estimate from either the Elastic Net or XGBoost model.

To estimate the relationship between birth weight and structural MRI measures, separate models were fitted using the following formula:

```
bw ~ bs(age, df=5) + vol_meas + C(site) + C(sex_female)
```

where `vol_meas` denotes the volumetric measure of interest.

Because longitudinal measurements are noisy over short time intervals, we used a estimate of measurement noise for each subject based on the time interval between the first and last session. Assuming that imaging sessions are approximately equally spaced over the observation period, the measurement noise is approximately proportional to the inverse squared observation time [10]:

$$Var[\alpha] \propto \frac{1}{\Delta t^2}. \tag{2}$$

For the longitudinal association analyses, we therefore weighted each subject by the squared total observation time. This ensures that slope estimates derived from longer observation windows contribute proportionally more to the regression estimates. This approach is broadly similar to excluding subjects with very short observation periods, but allows all available data to contribute to the analysis.

#### 2.4 Tau PET association

Estimated tau levels from PET were obtained from ADNI, Prevent-AD, DLBS, and HABS. We did not process the PET scans ourselves, but used the estimated values as provided by each contributing source. For some datasets, the temporal overlap between brain MRI and PET was limited, as shown in Supplementary Fig. 2.

Tau levels increase relatively predictably over time, making it possible to estimate tau levels at the time of MRI scanning. Since not all PET scans were acquired at the same visit as the structural MRI scans, we used Sample Iterative Local Approximation modelling, as described by [11], to estimate tau PET levels at each MRI scan time.

For each MRI session, tau positivity was defined using a threshold of 1.11 standardized uptake value ratio (SUVR) [12]. The same threshold was used across all cohorts.

Logistic regression was used to estimate the association between tau PET positivity and brain age or volumetric measures.

Tau accumulation is a hallmark biomarker of late-life neurodegenerative disease and reflects an age-related process. The tau PET analyses were therefore used to test whether brain age models were sensitive to individually varying age-related change. We compared the sensitivity of brain age gap with the sensitivity of the simpler volumetric measures.

For the association between tau positivity and brain age gap, separate logistic regression models were fitted for the Elastic Net and XGBoost brain age gaps using the following formula:

```
tau_pos ~ bs(age, df=3) + brain_age_gap + C(site) + C(sex_female)
```

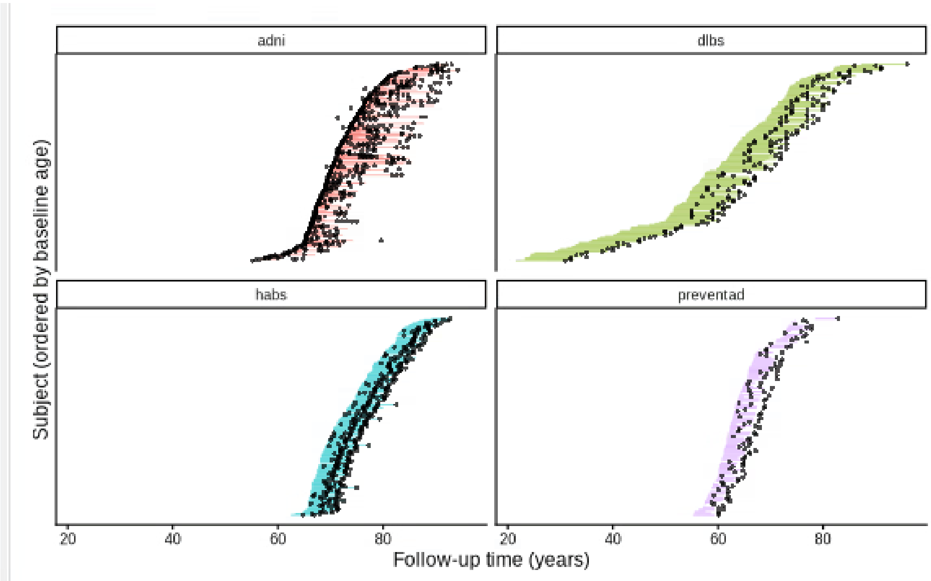


**Fig. 2** PET and MRI scans for the contributing datasets. Black dots indicate PET scans, while coloured dots indicate MRI scans.

For the association between tau positivity and volumetric measures, separate logistic regression models were fitted using the following formula:

```
tau_pos ~ bs(age, df=3) + vol_meas + C(site) + C(sex_female) + ICV
```

where `vol_meas` denotes the volumetric measure of interest. For the volumetric measures, the sign was flipped where necessary so that higher values consistently reflected a more adverse brain phenotype and therefore higher expected risk of tau PET positivity.

Similar to the birth weight analysis, longitudinal measures were weighted by the squared total observation time.

To compare Elastic Net brain age gap with selected volumetric measures directly, both predictors were included in the same logistic regression model. The volumetric measures tested were hippocampal volume, hippocampal volume change, total grey matter volume, and total grey matter volume change. For each comparison, the following logistic regression model was fitted:

```
tau_pos ~ age + vol_meas + elastic_net_gap
          + C(site) + C(sex_female) + ICV
```

where `vol_meas` denotes the volumetric measure being compared with Elastic Net brain age gap. For longitudinal volumetric measures, observations were weighted by the squared total observation time. Age, the volumetric measure, and Elastic Net brain age gap were standardized before model fitting. Two-sided Wald tests were then used to test whether the coefficient for the volumetric measure differed from the coefficient for Elastic Net brain age gap.

#### 2.5 Acknowledgements to data provides

Parts of the data collection and sharing for this project was funded by the Alzheimer's Disease Neuroimaging Initiative (ADNI) (National Institutes of Health Grant U01 AG024904) and DOD ADNI (Department of Defense award number W81XWH-12-2-0012). ADNI is funded by the National Institute on Aging, the National Institute of Biomedical Imaging and Bioengineering, and through generous contributions from the following: AbbVie, Alzheimer's Association; Alzheimer's Drug Discovery Foundation; Araclon Biotech; BioClinica, Inc.;Biogen; Bristol-Myers Squibb Company; CereSpir, Inc.; Cogstate; Eisai Inc.; Elan Pharmaceuticals, Inc.; Eli Lilly and Company; EuroImmun; F. Hoffmann-La Roche Ltd and its affiliated company Genentech, Inc.; Fujirebio; GE Healthcare; IXICO Ltd.; Janssen Alzheimer Immunotherapy Research & Development, LLC.; Johnson & Johnson Pharmaceutical Research & Development LLC.; Lumosity; Lundbeck; Merck & Co., Inc.; Meso Scale Diagnostics, LLC.; NeuroRx Research; Neurotrack Technologies; Novartis Pharmaceuticals Corporation; Pfizer Inc.; Piramal Imaging; Servier; Takeda Pharmaceutical Company; and Transition Therapeutics. The Canadian Institutes of Health Research is providing funds to support ADNI clinical sites in Canada. Private sector contributions are facilitated by the Foundation for the National Institutes of Health (www.fnih.org). The grantee organization is the Northern California Institute for Research and Education, and the study is coordinated by the Alzheimer's Therapeutic Research Institute at the University of Southern California. ADNI data are disseminated by the Laboratory for Neuro Imaging at the University of Southern California.

UK Biobank is generously supported by its founding funders the Wellcome Trust and UK Medical Research Council, as well as the Department of Health, Scottish Government, the Northwest Regional Development Agency, British Heart Foundation and Cancer Research UK. The organisation has over 150 dedicated members of staff, based in multiple locations across the UK.

Parts of the data used in preparation of this article were obtained from the Pre-Symptomatic Evaluation of Novel or Experimental Treatments for Alzheimer's Disease (Prevent AD) program.

Data used in the preparation of this article were also obtained in part from the Harvard Aging Brain Study (HABS - P01AG036694; https://habs.mgh.harvard.edu). The HABS study was launched in 2010, funded by the National Institute on Aging. and is led by principal investigators Reisa A. Sperling MD and Keith A. Johnson MD at Massachusetts General Hospital/Harvard Medical School in Boston, MA.

### References


[1] Mueller, S. G. *et al.* The Alzheimer's disease neuroimaging initiative. *Neuroimaging Clinics* **15**, 869–877 (2005).

[2] Petersen, R. *et al.* Alzheimer's disease Neuroimaging Initiative (ADNI) clinical characterization. *Neurology* **74**, 201–209 (2010).

[3] Tremblay-Mercier, J. *et al.* Open science datasets from PREVENT-AD, a longitudinal cohort of pre-symptomatic Alzheimer's disease. *NeuroImage: Clinical* **31**, 102733 (2021).

[4] Park, D. *et al.* The dallas lifespan brain study: A comprehensive adult lifespan data set of brain and cognitive aging. *Scientific Data* **12** (2025).

[5] Dagley, A. *et al.* Harvard aging brain study: dataset and accessibility. *NeuroImage* **144**, 255–258 (2017).

[6] Miller, K. *et al.* Multimodal population brain imaging in the UK Biobank prospective epidemiological study. *Nature Neuroscience* **19**, 1523–1536 (2016).

[7] Walhovd, K. *et al.* Neurodevelopmental origins of lifespan changes in brain and cognition. *Proceedings of the National Academy of Sciences* **113**, 9357–9362 (2016).

[8] Walhovd, K. B. *et al.* Healthy minds 0–100 years: Optimising the use of european brain imaging cohorts ("lifebrain"). *European Psychiatry* **50**, 47–56 (2018).

[9] Smith, S. M., Vidaurre, D., Alfaro-Almagro, F., Nichols, T. E. & Miller, K. L. Estimation of brain age delta from brain imaging. *Neuroimage* **200**, 528–539 (2019).

[10] Grødem, E. O. *et al.* Stable individual differences dominate adult brain volume variation until later life. *Imaging Neuroscience* (2026).

[11] Betthauser, T. J. *et al.* Multi-method investigation of factors influencing amyloid onset and impairment in three cohorts. *Brain* **145**, 4065–4079 (2022). URL https://doi.org/10.1093/brain/awac213.

[12] Maass, A. *et al.* Comparison of multiple tau-PET measures as biomarkers in aging and Alzheimer's disease. *NeuroImage* **157**, 448–463 (2017).
